# The use of wearable technology to objectively measure sleep quality and physical activity among pregnant women in urban Lima, Peru: A pilot feasibility study

**DOI:** 10.1101/2019.12.29.19016097

**Authors:** Jerome T. Galea, Karen Ramos, Julia Coit, Lauren E. Friedman, Carmen Contreras, Milagros Dueñas, Giuliana Noris Hernandez, Caroline Muster, Leonid Lecca, Bizu Gelaye

**Author notes:** **Corresponding author:** Jerome Galea, Ph.D., M.S.W., Assistant Professor, School of Social Work, University of South Florida, 4202 E. Fowler Avenue, MHC 1420, Tampa, FL 33620. Jerome Galea and Karen Ramos contributed equally to this manuscript. &.

## Abstract

Sleep quality and physical activity can affect the mental and physical health of pregnant women and their unborn babies. We investigated the feasibility of assessing sleep quality and physical activity among pregnant women in Peru. Twenty women maintained sleep logs and wore ActiSleep devices for seven consecutive days; 13 had sufficient data for analysis. Mean sleep duration was 6.9 hours (SD = 1.4). Sleep efficiency was 77.9%. Participants averaged of 6,029 steps per day (SD = 3,087). Objective assessment of sleep quality and physical activity was feasible. Wearable technology has applications in healthcare to improve sleep quality and physical activity.

## Introduction

Pregnant women are at increased risk for sleep disturbances^1,2^, poor sleep quality^3^, insomnia^4^, and changes in sleep architecture, the cyclical pattern of sleep as it shifts between different stages^5^. These changes are associated with increased prevalence of mood disorders, including depression^6^, suicidal ideation, and anxiety^7^. Sleep quality, which deteriorates during pregnancy^8^, is also associated with adverse postnatal outcomes including pre-eclampsia, hypertension, low birth weight, prematurity, and stillbirth^9,10,11^. Lower physical activity among pregnant women is associated with multiple health risks including obesity, gestational diabetes, Type-2 diabetes, cardiovascular disease, and cesarean delivery^12,13^.

Accordingly, sleep quality and physical activity are key intervention targets for improving pregnant women’s health and pregnancy outcomes. Relatively few methods exist, however, to evaluate sleep and activity outside of clinical settings. For example, though polysomnography is the gold standard for evaluating sleep, it can only be conducted in hospitals or sleep centers. In contrast, though lacking the precision of inpatient testing, wearable devices offer an inexpensive way to gather basic data on sleep and physical activity. In resource-constrained settings, wearable devices may be the only way to objectively collect such data. Poudyal and colleagues^14^ proposed the use of wearable sensors to identify the risk of postpartum depression for adolescent mothers in Nepal. However, the feasibility of using such devices in resource-constrained settings among women during pregnancy is unknown. The objective of this study was to investigate the feasibility of assessing sleep quality and physical activity among pregnant women in Peru.

## Methods

Participants resided in an impoverished, peri-urban district in northern Lima that with limited access to health services. The international nonprofit health organization Partners in Health (locally Socios En Salud) operates several health-related services in the area, including a maternal health program (project “SAMI”), from which participants were recruited.

Participants wore ActiSleep monitors (ActiLife, ActiGraph R&D, Florida, USA) on their nondominant wrist for seven consecutive days. They also maintained daily sleep logs, noting the times they got into bed at night, arose in the morning, and the start and end times of any naps. Sociodemographic and pregnancy data were abstracted from the SAMI program records.

All participants provided written informed consent prior to beginning the study. The study was approved by the institutional review boards of the Cayetano Heredia University in Lima, Peru (#17012) and the Harvard T.H. Chan School of Public Health in Boston, MA. No compensation was provided to participants.

Actigraphy sleep data were collected using ActiSleep monitors and analyzed using ActiLife 6 software. ActiSleep monitors are similar to large digital wristwatches and provide estimates of sleep onset, sleep latency, wake after sleep onset (WASO), number and length of awakenings, sleep duration, and sleep efficiency. Sleep latency is the length of time it takes to fall asleep, calculated as the time between ‘lights off’ to the first 3 consecutive minutes of sleep. WASO refers to the number of minutes awake between sleep onset and time of final waking.

Sleep efficiency is the proportion of the estimated sleep periods spent asleep. Daily step counts were recorded for each participant as measurements of physical activity.

ActiSleep monitors collect data in one-minute intervals, using zero-crossing modes and the Sadeh sleep algorithm. Means were calculated for each nighttime sleep parameter and described using standard deviations (SD), or medians and corresponding interquartile ranges (IQR) for non-parametric variables. Categorical variables were described using number and percentages (%).

## Results

Half of the participants (N=10) had complete seven-day ActiSleep sleep data. One participant had six days of data and two participants had four days of data, for a total of N = 13 participants included in the analysis. The remaining (N=7) participants were not included, due to missing data.

Average gestational age at time of interview was 22 weeks (SD = 4.9). Four women were nulliparous. Mean age was 26.3 years (SD=3.9). Average weight in early pregnancy was 63 kg (SD=7.2). Sleep and physical activity parameters are presented in Table 1. The median time of day for sleep onset was 21:15. Mean sleep latency was 17.3 minutes (SD = 18.8). WASO was 116 minutes (SD = 63.5). Mean sleep duration was 6.9 hours (SD = 1.4). Mean number of awakenings was 20.4 (SD = 6.7). Sleep efficiency was 77.9%. Participants had an average of 6,029 steps per day (SD = 3,087) (Figure 1), which falls within the range of ‘physically inactive’^13^.

**Table 1:** Sleep and physical activity parameters for participants in pilot study

| Parameter | Total Participants (N = 13) |
| --- | --- |
| | Mean ± SD |
| Average step counts | 6,029 ± 3,087 |
| Steps per minute | 3.4 ± 1.9 |
| Lux average counts | 5.7 ± 4.3 |
| Maximum lux value | 1640 ± 882 |
| Sleep onset (median ± IQR) | 21:15 [13:44 – 22:42] |
| Sleep latency (minutes) | 17.3 ± 18.8 |
| WASO (minutes) | 116 ± 63.5 |
| Number of awakenings (n) | 20.4 ± 6.7 |
| Awakening length (minutes) | 7.7 ± 5.8 |
| Sleep duration (hours) | 6.9 ± 1.4 |
| Nap sleep duration (hours) (N = 10) | 1.0 ± 0.7 |
| Sleep efficiency (%) | 77.9 ± 9.6 |

**Notes:** Actigraphy data were collected in one-minute epochs using the zero-crossing modes; and the ‘Sadeh’ sleep algorithm was used. Each observation was treated as a separate measure for the preliminary sleep parameter analyses. Data from each participant are highly correlated, which must be considered in subsequent analyses.

**Figure 1:**
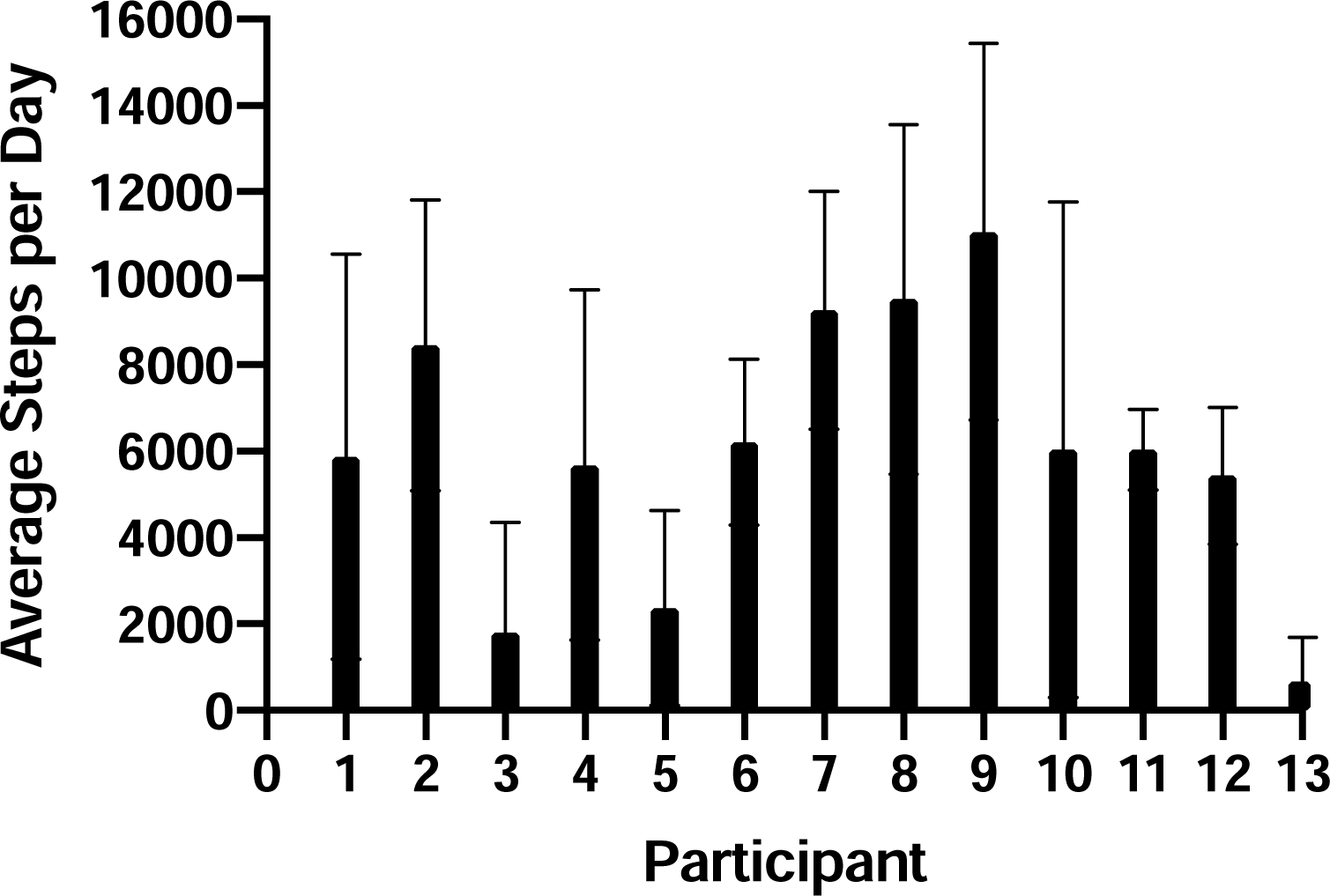
Average step counts for participants in pilot study (N = 13)

## Discussion

Using wearable technology to measure sleep quality and physical activity among pregnant women in a resource-constrained setting in urban Lima was somewhat feasible. However, 35% of participants collected less than four days of data. Although we could not confirm why participants did not use the ActiSleep monitor during the entire seven days, study staff reported anecdotally that some women removed the device during the day due to discomfort or to prevent exposure to water (e.g., from bathing). Once removed, some women forgot to put the monitor back on in the evening.

Because this feasibility pilot was embedded in a larger intervention with separate objectives, our teams were limited in our ability to provide additional support beyond a basic tutorial during data collection. Future studies using wearable technology in these populations should include a qualitative component to assess the factors related to acceptability of the device. Kohrt and colleagues^15^ recently proposed a qualitative methodology called Cultural Assessment of Passive Data collection Technology (QualCAPDT) to identify cultural norms and family preferences affecting the uptake of new technologies, like wearable devices, among children and their caregivers in Nepal. Application of QualCAPDT or similar programs could help explain why >1/3 of participants in this study did not consistently adhere to the wearable device protocol.

Wearable devices represent relatively untapped technology for healthcare interventions in resource-constrained environments. There is demonstrated promise in their potential to provide valuable information to inform treatment of mental and physical illness. This proof of concept study demonstrates that, even with limited instruction and support, it is possible to collect sleep and physical activity data on pregnant women in a resource-constrained environment.

## Data Availability

Data available upon request.

## Declarations

### Authors’ contributions

JTG, LEF, and BG designed the study. All authors implemented the study. JTG, JC, LEF, and BG analyzed and interpreted the data. JTG, KR, JC, LEF, CM, and BG wrote the manuscript. All authors read and approved the final manuscript.

## Acknowledgements

The authors thank the women who participated in the study.

## Ethical approval

The study was approved by the institutional review boards of the Cayetano Heredia University in Lima, Peru (#17012) and the Harvard T.H. Chan School of Public Health in Boston, MA.

